# PHARMACEUTICAL CARE INTERVENTIONS OF INPATIENT PRESCRIPTIONS AT HOSPITAL PAKAR SULTANAH FATIMAH MUAR

**DOI:** 10.1101/2025.03.10.25323704

**Authors:** Rohidayah Abd Majid, Nur Atika Abdul Manap, Farizal Farid Basheer, Rohaniza Yahaya, Nurul Hanis Mohd Sabari

## Abstract

**Introduction:** Prescribing medications plays a vital role in patient healthcare. The rational drug use is now a significant concern for public health due to incorrect medication prescribing. In promoting rational evidence-based prescribing, prescriptions will be screened and reviewed by pharmacists before medications are dispensed whereby pharmaceutical care intervention (PCI) will be conducted.

**Methods:** This study aimed to evaluate the prevalence and types of PCI detected at an inpatient pharmacy and to identify the stage of dispensing process where the PCI are frequently detected. The PCI that included was focusing on the prescribing errors. A cross-sectional observational study was conducted over a period of three months started from 1st March 2023 until 31st May 2023 where new medication orders using Pharmacy Information System (PhIS) of all patients warded screened by Inpatient Pharmacy Department were included.

**Results:** The prevalence of PCI was 0.006%. The most common type of interventions performed were the prescribed frequency (31.5%) followed by dose (30.0%), drug (19.2%) and polypharmacy (10.8%). The drug category based on ATC classification with a high percentage of interventions was anti-infective for systemic use (34.6%) followed by nervous system (16.2%) and alimentary tract and metabolism (14.7%). Half of the PCIs were detected during screening stage (51.2%) whiles 36.0% were detected during counterchecking and the least detected is during medication filling (12.8%). The highest prescribing errors was from medical wards (50.7%), followed by surgical (24.1%) and orthopaedics (14.8%).

**Conclusion:** The prescribers and clinical pharmacist and inpatient pharmacist are doing well in maintaining patient care. Prescriptions that involve drug category of anti-infective required more attentions especially on drug choice, dose, frequency, and polypharmacy.

## Introduction

To enhance rational and evidence-based prescribing, pharmacists will screen and review prescriptions before medications are dispensed whereby pharmaceutical care intervention (PCI) will be conducted on medication errors. Unsafe medication practices and medication errors constitute a primary cause of injury and preventable harm in healthcare systems worldwide. Because of incorrect medication prescribing, rational drug usage has become an important public health issue[1-3]. World Health Organization (WHO) has estimated that more than 50% of all medicines are prescribed, dispensed, inappropriately across the world and 50% of patients did not take them properly [3].

Medication-related problems frequently arise in hospital wards, occurring during various stages such as prescribing, transcribing, dispensing, administering, adherence to, or monitoring of drugs [4]. A prospective observational study of 681 patients by Elhabil MK *et al* showed that 221 medication errors (ME) occurred in 29.22% of patients and majority of the ME was prescribing errors (82.8%) [2]. Prescribing errors (PEs) are defined as “a clinically meaningful prescribing error that occurs as a result of a prescribing decision or the prescription writing process resulting in an unintentional significant reduction in the probability of treatment being timely and effective or in increasing the risk of harm when compared to generally accepted practice” [5]. An effective PCI by the pharmacist to adjust patient medication or therapy is crucial not only for reducing the PEs and unnecessary costs associated with adverse drug events but also for optimizing pharmacotherapy outcomes [6].

PEs are a significant concern across both community practices and hospitals due to their potential to cause harm to patients. Published studies have demonstrated that PE can arise from prescribing incorrect medications or dosages, incorrect frequency, duplicate orders, and the prescription of medications that are restricted[5, 7-9]. Alzahrani et al reported that wrong dose (54.3 %) and unauthorized prescription (21.9 %) were the most encountered PEs while the highest observed PCI to prevent PEs were related to dose adjustments (44.0 %), restricted medication approvals (21.9 %), and therapeutic duplications (11 %) [5].

Hospital admission presents an ideal opportunity for pharmacists to conduct a thorough review of a patient’s pharmacotherapy. This is particularly valuable in cases where hospitalization may be related to medication issues or in patients with complex medication schedules, such as elderly individuals with multiple medical conditions who are receiving multiple medications [6]. However, in hospital settings, handwritten prescriptions often contain issues that must be addressed by ward or inpatient pharmacists. To expedite the dispensing process particularly when medications are urgently needed for critically ill patients, a fool-proof and reliable computerized system could serve as an alternative to replace manual order entries by physicians. A systematic review and meta-analysis by Gates PJ et al concluded that the computerizes system able to reduce prescribing error rates [10].

Hospital Pakar Sultanah Fatimah, a public hospital in the southernmost state of Peninsular Malaysia has incorporated the Pharmacy Information System (PhIS) as an e-prescription system to enhance and thereby lower the chance of prescribing errors. Nonetheless, there is a lack of studies reviewing the implementation of an e-prescribing system within inpatient pharmacies in local settings. Therefore, this study aimed to evaluate the prevalence and types of PCI detected at an inpatient pharmacy and to identify the stage of dispensing process where the PCI are frequently detected.

## Methodology

### Design and Setting

A cross-sectional prospective study was conducted over a period of three months started from 1st March 2023 until 31st May 2023 in the main inpatient (ward supply) pharmacy department of Hospital Pakar Sultanah Fatimah, Muar, Johor, Malaysia. This is a district government hospital that serves around 500 beds. The ward supply pharmacy receives and screens around 9000 to 12000 new inpatient prescriptions daily. The medications were prescribed using online system which is PhIS where the eprescription will be screened by trained pharmacists before proceeding with filling and then counterchecking before dispensing the medication to wards.

### Data Collection and Analysis

All e-prescriptions screened from 8.00 am to 5.00 pm during the study period excluding weekend and public holiday were included in this study. The PCI were detected during the screening, filling, and counterchecking stages which then prescribers were contacted for the intervention. This study aimed to evaluate the prevalence and types of PCI detected and the common drugs involved in the PCI at the ward supply pharmacy and to identify the stage of dispensing process where the PCI are frequently detected. The PCIs were recorded in a prescription intervention form that provided data on ward specialty, interventions by pharmacist, category of drug involved and dispensing stage where the PCI detected. Data confidentiality was preserved, including the identities of the intervening pharmacist, the prescriber, and the patient. All data were transcribed and analysed using Microsoft Excel 2023. Descriptive statistical analysis was used to assess the rate of interventions, types of prescription interventions, and categories of drugs involved. The study was conducted according to the guidelines of the Declaration of Helsinki, and approved by the Medical Research Ethics Committee, Ministry of Health Malaysia (Ref: 23-02942-XL6 (2)).

## Result

### Prevalence of PCI

A total of 33223 new inpatient e-prescriptions were received and screened by the ward supply pharmacy from different specialty wards during the 3 months study period. Out of the total screened prescription, there were 203 PCI detected which produce 0.006% rate of PCI. The most common type of interventions recorded were the prescribed frequency (31.5%) followed by dose (30.0%), drug (19.2%) and polypharmacy (10.8%) as shown in Table 1. The highest prescribing errors was from medical wards (50.7%), followed by surgical (24.1%) and orthopaedics (14.8%) (Table 2). The distribution of types of interventions in different ward specialty is illustrated in Figure 1. When comparing the types of interventions at different ward specialty, result from this study reported that more than half of the PCI for drug choice (59.05%), frequency (54.7%) and polypharmacy (54.6%) were from medical wards. Even though number of PCI for duration is only 10, however 70% of them were from medical wards. Medical and surgical wards contributed about the same percentage of PCI for dose, 39.3% and 32.8% respectively. Whiles, prescriptions from neonatal intensive care unit were only intervened for dose (4.9%) and frequency (1.6%).

**Table 1:** Types of PCI in Different Ward Specialty.

| Types of Intervention | N=203<br>n(%) | Ward Specialty |  |  |  |  |  |
| --- | --- | --- | --- | --- | --- | --- | --- |
|  |  | Medical<br>n(%) | Surgical<br>n(%) | Orthopedics<br>n(%) | Intensive<br>care<br>n(%) | Neonatal<br>intensive<br>care<br>n(%) | Others<br>n(%) |
| Drug | 39 (19.2) | 23(59.0) | 8(20.5) | 4(10.3) | 4(10.3) | - | - |
| Dose | 61 (30.0) | 24(39.3) | 20(32.8) | 8(13.1) | 5(8.2) | 3(4.9) | 1.6 |
| Frequency | 64 (31.5) | 35(54.7) | 12(18.8) | 13(20.3) | 1(1.6) | 1(1.6) | 3.1 |
| Duration | 10 (4.9) | 7(70.0) | 1(10.0) | - | 1(10.0) | - | 10.0 |
| Polypharmacy | 22 (10.8) | 12(54.6) | 5(22.7) | 3(13.6) | 2(9.1) | - | - |
| Others | 7 (3.4) | 2(28.6) | 3(42.9) | 2(28.6) | - | - | - |

**Table 2:** Distribution of PCI in Different Ward Specialty.

| Ward Specialty | Number N=203<br>n(%) |
| --- | --- |
| Medical | 103(50.7%) |
| Surgical | 49(24.1%) |
| Orthopedics | 30(14.8%) |
| Intensive care | 13(6.4%) |
| Neonatal intensive care | 4(2.0%) |
| Others | 4(2.0%) |

The above data are pictured in the next graph

**Figure 1.**
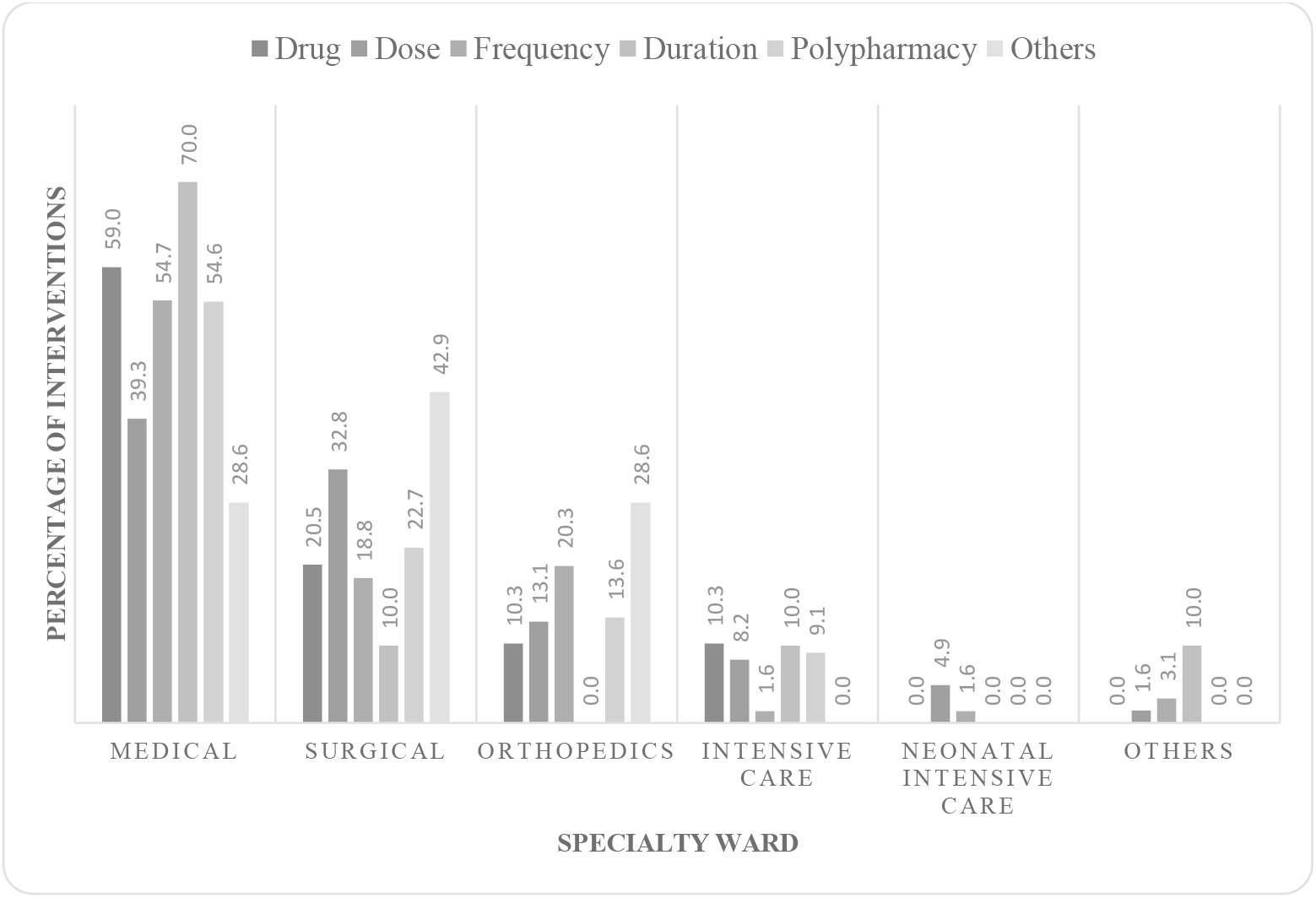
Types of Intervention in Different Ward Specialty

### Categories of Drugs Involved in the PCI

Table 3 shows that overall, the most common drugs being intervened were from anti-infective for systemic use group (34.6%) followed by nervous system (16.2%) and alimentary tract and metabolism (14.7%). Drug from group genito unirary system and sex hormones is the least intervened drug.

**Table 3:** Distribution of PCI Based on Drug Category.

| Categories of drug | Total numbers (N) | Percentage (%) |
| --- | --- | --- |
| Anti-infective for systemic use | 66 | 34.6 |
| Nervous system | 31 | 16.2 |
| Alimentary tract and metabolism | 28 | 14.7 |
| Cardiovascular system | 23 | 12.0 |
| Respiratory system | 12 | 6.3 |
| Blood & blood forming organs | 8 | 4.2 |
| Musculoskeletal system | 8 | 4.2 |
| Various | 7 | 3.7 |
| Systemic hormonal preparations | 6 | 3.1 |
| Genito urinary system and sex hormones | 2 | 1.0 |

When comparing the drug categories that commonly intervened in different ward specialty as depicted in Table 4, we found out that anti-infectives for systemic use were the most frequently intervened drug from medical (31.3%), surgical (37.8%), orthopaedic (42.9%) and intensive care wards (33.3%). From the total interventions performed in orthopaedic ward, nervous system drugs were more frequently intervened (28.6%) compared to medical wards (17.2%) and surgical wards (13.3%). Whiles, surgical wards were more frequently intervened for alimentary tract and metabolism drugs (22.2%) after anti-infectives for systemic use drugs. Likewise, intensive care wards were more frequently intervened for respiratory system drugs (25.0%).

**Table 4:** Categories of Drugs Intervened in Different Ward Specialty.

| **Drug Categories (%)** | **Medical** | **Surgical** | **Orthopaedics** | **Intensive care** | **Neonatal intensive care** | **Others** |
| --- | --- | --- | --- | --- | --- | --- |
| Anti-infectives for systemic use | 31.3 | 37.8 | 42.9 | 33.3 | 25.0 | 33.3 |
| Cardiovascular system | 17.2 | 6.7 | 10.7 | - | - | - |
| Nervous system | 17.2 | 13.3 | 28.6 | - | - | - |
| Alimentary tract and metabolism | 13.1 | 22.2 | 3.6 | 16.7 | 25.0 | 33.3 |
| Respiratory system | 7.1 | - | - | 25.0 | 25.0 | 33.3 |
| Systemic hormonal preparations | 4.0 | 4.4 | - | - | - | - |
| Blood and blood forming organs | 3.0 | 4.4 | 3.6 | 16.7 | - | - |
| Musculoskeletal system | 3.0 | 6.7 | 7.1 | - | - | - |
| Genito urinary system and sex hormones | - | 4.4 | - | - | - | - |
| Various | 4.0 | - | 3.6 | 8.3 | 25.0 | - |

### PCI at Different Stage of Dispensing

Table 5 describes the frequency of PCI detected at different stage of dispensing which were screening, filling, and counterchecking. Half of the interventions (51.2%) were detected during screening stage where PCI for dose was mostly intervened. Most of the wrong frequency were detected during counterchecking stage (39.7%).

**Table 5:** Frequency of PCI Detected at Different Stage of Dispensing

| Type of Intervention | Intervention by stage of dispensing process, n (%) |  |  |
| --- | --- | --- | --- |
|  | Screening<br>N= 104(51.2) | Filling<br>N= 26(12.8) | Counterchecking<br>N= 73(36.0) |
| Drug | 19 (18.3) | 7(26.9) | 13(17.8) |
| Dose | 44(42.3) | 5(19.2) | 12(16.4) |
| Frequency | 28(26.9) | 7(26.9) | 29(39.7) |
| Duration | 5(4.8) | 1(3.8) | 4(5.5) |
| Polypharmacy | 5(4.8) | 4(15.4) | 13(17.8) |
| Others | 3(3.0) | 2(7.7) | 2(2.7) |

## Discussion

Rate of PCI per prescription screened by pharmacists in the main inpatient (ward supply) pharmacy (0.006%) was very low compared to a study done to evaluate the intervention of e-prescription in inpatient setting at Malaysian Public Hospital(3.2%) by Ooi P. L. et al[7] and a study done by Safaie etal on intervention in Postoperative Cardiac Intensive Care Unit (0.19%) [11]. E-prescribing has been promoted as a potential mechanism for reducing medication errors, supported by studies demonstrating lower rates of interventions [7, 12]. The e-prescription can significantly reduce prescribing errors by automating the process of prescription transmission and order entry. This study shows that the prescribers and inpatient pharmacists were doing well in maintaining patient care. Besides, the PCI detected in the inpatient pharmacy was very low highly due to the role of ward pharmacists especially in the medical wards. Ward pharmacists work closely with other health professionals to ensure that each patient receives the correct medication throughout their hospital stay. Their involvement is vital in safeguarding against medication errors during patient admission, hospitalization, and discharge[2].

Frequency, dose, drug, and polypharmacy are the main encountered type of interventions in this study. The same trend was also reported in several other studies [5, 7, 9, 13]. Wrong or inappropriate frequency and dose were the highest type of PCI recorded with about the same percentage. A study done by Alzahrani et al on pharmacist interventions related to reported prescribing errors exhibited that half (54.3%) of the interventions were on the wrong dose however, wrong frequency was only accounted with 2.3% [5] . Whiles, Ooi P.L et al reported that wrong frequency (13.7%) was the third common type of interventions performed [7]. A systematic review of 9 studies done by Alazani et al. extracted that dosage problems or wrong dosage was the most frequent type of prescribing errors detected in the studies with the highest percentage in 33% (3/9) of the studies [14]. Dose interventions involve modifications to the amount of medication a patient receives per administration. These interventions are crucial for ensuring the correct therapeutic dose is given, which varies based on factors like age, weight, kidney function, and other patient-specific factors. Frequency interventions, which involve adjusting how often a medication is administered, are indeed common in inpatient settings. These interventions are critical for ensuring that medications are given at the correct intervals to maximize efficacy and minimize the risk of side effects or toxicity [15].

Half of the PCI for wrong drug, frequency and polypharmacy were from the medical wards. Medical wards also contribute the highest intervention for duration compared to others. Alzahrani et al also presented that wrong dose was more common detected from medical wards which consistent with our findings [5]. In this study, surgical wards were the second department after medical that frequently intervened by the inpatient pharmacy. Whereas, Ooi P.L et al reported that the greatest number of interventions was from surgical wards (11.7%)[7]. Medical wards have higher prevalence of prescribing errors can be due to the complexity of cases, frequent medication changes and dose modifications especially for unstable patients and patients with multiple comorbidities.

We found that most of the drug categories being intervened were from anti-infective for systemic use group. This result is consistent with Ooi P.L et al with 33.8% of the intervention performed was from anti-infective agents. Alzahrani et al also produced the same result with 49.2% of the prescribing errors were related to anti-infective for systemic use group. Our finding was also concurring with other studies [5, 7, 9, 11, 13]. Anti-infectives for systemic use were the most frequently intervened drug from medical, surgical, orthopaedic, and intensive care wards. This shows that antibiotics are indeed among the most frequently intervened drug groups in inpatient settings. The reason for this might be explained as the drug choice, dosage, and frequency of the anti-infective drugs especially for systemic use have varies adjustment which lead to more interventions required. The interventions are often focus on ensuring appropriate dosing, frequency, and selection, as well as preventing overuse or misuse that could contribute to antibiotic resistance[16].

This study showed that half of the interventions were detected during screening stage. The screening of medication orders is usually conducted by senior pharmacist in the inpatient pharmacy which led to more PCI detected at this stage. This stage involves reviewing prescriptions for appropriateness before filling, followed by screening for the prescribing errors during the filling and counterchecking stages [10].

## Limitation

The study was conducted at only one healthcare centre, limiting the generalizability of the results to other settings with different patient populations or healthcare practices. The PCIs were identified based solely on prescription records, without considering potential clinical judgments that were not documented. This could lead to an incomplete assessment of prescribing errors. Therefore, future research should incorporate a multicentre design and involve physicians in the assessment of prescribing errors.

## Conclusion

The prescribers and clinical pharmacist and inpatient pharmacist are doing well in maintaining patient care. Prescriptions that involve drug category of anti-infective required more attentions especially on drug choice, dose, frequency, and polypharmacy. Most of the PCIs were successfully detected at screening stage and this is essential for optimizing medication use and decrease drug related problem before dispensing.

## Data Availability

All data produced in the present study are available upon reasonable request to the authors

## Acknowledgement

We would like to thank the Director General of Health, Malaysia, for his permission to publish the findings from this study. We also would like to thank the head and staff of the Inpatient Pharmacy Department of the Hospital Pakar Sultanah Fatimah for the permission and assisting us in the data collection.

## Notes

### Competing Interest Statement

The authors have declared no competing interest.

### Funding Statement

This study did not receive any funding

### Author Declarations

The study was conducted according to the guidelines of the Declaration of Helsinki, and approved by the Medical Research Ethics Committee, Ministry of Health Malaysia (Ref: 23-02942-XL6 (2)).

## References

1. Challenge, W.G.P.S., World health organization Global patient safety challenge: Medication Without Harm. 2017, World Health Organization 2017: WHO/HIS/SDS/2017.6. p. 1-16.

2. Elhabil, MK, et al., Impact of Clinical Pharmacist-Led Interventions on Drug-Related Problems Among Pediatric Cardiology Patients: First Palestinian Experience. Integrated pharmacy research & practice, 2022. 11.

3. Organization, W.H., Promoting rational use of medicines: core components. 2002, World Health Organization.

4. Gebre, M., et al., Medication Errors Among Hospitalized Adults in Medical Wards of Nekemte Specialized Hospital, West Ethiopia: A Prospective Observational Study. Drug, Healthcare and Patient Safety, 2021. 13: p. 221–228.

5. Alzahrani, A.A., et al., Description of pharmacists’ reported interventions to prevent prescribing errors among in hospital inpatients: a cross sectional retrospective study. BMC Health Services Research, 2021. 21(1): p. 432.

6. Dalton, K. and S. Byrne, Role of the pharmacist in reducing healthcare costs: current insights. Integr Pharm Res Pract, 2017. 6: p. 37–46.

7. Poh Ling, O., et al., Pharmacists’ Interventions on Electronic Prescriptions from Various Specialty Wards in a Malaysian Public Hospital: A Cross-Sectional Study. Pharmacy (Basel, Switzerland), 2021. 9(4).

8. Reckmann, M.H., et al., Does computerized provider order entry reduce prescribing errors for hospital inpatients? A systematic review. J Am Med Inform Assoc, 2009. 16(5): p. 613–23.

9. Anzan, M., et al., Prescribing errors and associated factors in discharge prescriptions in the emergency department: A prospective cross-sectional study. PLOS ONE, 2021. 16(1): p. e0245321.

10. Gates, P.J., et al., How effective are electronic medication systems in reducing medication error rates and associated harm among hospital inpatients? A systematic review and meta-analysis. J Am Med Inform Assoc, 2021. 28(1): p. 167–176.

11. Safaie, N., H. Azizi, and S. Khiali, The Impact of Clinical Pharmacist Interventions on Medication Errors Management in the Postoperative Cardiac Intensive Care Unit. Pharmaceutical Sciences, 2021. 27.

12. Osmani, F., et al., Evaluation of the effectiveness of electronic prescription in reducing medical and medical errors (systematic review study). Ann Pharm Fr, 2023. 81(3): p. 433–445.

13. Albayrak, A., et al., Clinical pharmacist assessment of drug-related problems among intensive care unit patients in a Turkish university hospital. BMC health services research, 2022. 22(1).

14. Alanazi, M.A., M.P. Tully, and P.J. Lewis, A systematic review of the prevalence and incidence of prescribing errors with high-risk medicines in hospitals. Journal of Clinical Pharmacy and Therapeutics, 2016. 41(3): p. 239–245.

15. Robert, S.A., et al., An Evaluation of Interventions by Clinical Pharmacists in a Tertiary Hospital. Malaysian Journal of Pharmacy (MJP), 2021. 7(2): p. 3–6.

16. Jantarathaneewat, K., B. Camins, and A. Apisarnthanarak, The role of the clinical pharmacist in antimicrobial stewardship in Asia: A review. Antimicrob Steward Healthc Epidemiol, 2022. 2(1): p. e176.

